# Factors affecting contact tracing for the early detection of Leprosy cases: A qualitative study in Ho, Volta Region, Ghana

**DOI:** 10.1101/2024.05.08.24306976

**Authors:** Araba Aseye Ahiabu, Philip Teg-Neefah Tabong

## Abstract

**Introduction:** Despite Ghana being at the elimination phase of Leprosy, new cases of Leprosy are recorded in the country annually. These cases are usually detected in the advanced stages when Grade 2 disabilities (G2D) have set in. The WHO regards high G2D rates as an indication that measures need to be taken especially with regard to early detection. Late detection of cases raises concern about the spread of the infection. Contact tracing of index cases of Leprosy is therefore key in containing the spread. If the current situation is left unattended, Ghana faces the challenge of retrogressing in its fight against Leprosy. The general objective is to explore the factors affecting contact tracing for early case detection of Leprosy.

**Methods:** This study employed a qualitative research methodology. Specifically, the study adopted the phenomenology study design. Maximum variation purposive sampling technique was used for people affected by Leprosy who were either on treatment at the Ho Polyclinic or had completed treatment, health workers of the Polyclinic and community members at Ho, in the Volta Region of Ghana. Data was collected using in-depth interviews and focus group discussions among selected participants. All IDIs and FGDs were audio-recorded and transcribed verbatim. The data was coded using NVivO version 14 and analysed thematically.

**Results:** The results of this study showed that a majority of the community members and people affected by Leprosy perceived that Leprosy is caused by supernatural forces and hence the disease is not transmissible. Factors such as the stigma associated with the disease, reliance on divine forms of healing, use of herbs and poor symptom recognition hinder early detection. The study also revealed gaps in contact tracing efforts such as the inadequacy of funds for contact tracing as well as the reluctance of some contacts to be involved in the process.

**Conclusion:** The study concluded that both community and health system factors affect early detection of Leprosy cases in Ho. This has the potential to undermine Ghana’s quest to eliminate Leprosy. There is a need to increase awareness of the importance of contact tracing for early detection of Leprosy.

## Introduction

Leprosy, also known as Hansen’s disease, is one of the oldest recorded diseases. It is an infectious disease caused by the bacteria, *Mycobacterium leprae* (1–3). The bacterium has a lengthy incubation period of up to 20 years (4). The onset of the disease is characterized by small lesions on the skin as well as peripheral neuropathies (1,2). If left untreated, Leprosy can lead to unaesthetic hyperpigmented patches of the skin, deformities, blindness, and cause damage to the peripheral nerves and the upper respiratory tract (5,6). Owing to this, early detection is key to mitigating its effects.

Huge strides have been made in the effort to combat the disease. Leprosy is treated with Multidrug Therapy (MDT) which replaced Dapsone monotherapy (4). The combination of Dapsone, Rifampicin, and Clofazimine has proven to be highly effective in treating Leprosy over the years (4). Despite the remarkable efficacy of the MDT, Leprosy is still being transmitted globally with a plateau of about 200,000 new cases annually, especially among low-income countries(7). The prevalence of Leprosy in low-income countries (LMICs) places it in the category of neglected tropical diseases (NTDs). Leprosy and other NTDs cost developing countries huge sums of money due to direct health costs, disability-adjusted years, reduced levels of educational attainment and stunted socio-economic growth (8,9).

The onset of Grade 2 Disabilities (G2D) is usually the marker for late detection. These are the physical deformities and neural impairments associated with Leprosy (6,10). As a sequel to this, the WHO regards high G2D rates as an indication that measures need to be taken especially concerning early case detection (World Health Organization, 2020). Some countries in the post-elimination phase still face challenges concerning the early detection of cases. Challenges with early detection compromise the effectiveness of contact tracing and hence has implications for the spread of the disease. Due to the lengthy incubation period, a person infected with Leprosy has the potential to infect many others while asymptomatic.

Contact tracing has proven to be an effective way to control the spread of the disease. Close contact with people living with the condition is the main mode of transmission of Leprosy (Hacker et al., 2021; Moet et al., 2006). Once an index case is detected, contacts of the affected individual are supposed to be traced and administered post-exposure prophylaxis (PEP) which is a single dose of Rifampicin (SDR). This prophylactic treatment has proven to be effective in reducing the occurrence of infection among contacts (13). The WHO recommends tracing household members of the index case as well as 20-25 social contacts and the administration of SDR to mitigate the spread of the disease (World Health Organization, 2020).

The Global Leprosy Strategy postulates that there should be the implementation of zero Leprosy road maps in all Leprosy endemic countries, scaling-up of Leprosy prevention and early detection measures, management of Leprosy and its related impairments as well as the eradication of the stigma associated with the disease while ensuring that human rights are upheld (World Health Organization, 2016).

Ghana is in the elimination phase of Leprosy. To attain this level, a nation is expected to report less than 1 new case per 10,000 population (14). New cases of Leprosy are recorded annually. These cases are detected in the later stages when Grade 2 disabilities (G2D) have set in. This situation certainly does not follow the WHO strategy for reducing Leprosy (14). Late detection not only has implications for disabilities but also for spreading the infection as Leprosy has a lengthy incubation period of up to 20 years without symptoms (15).

As of September 2022, Ghana had recorded 221 cases of Leprosy across the country with a majority of cases reported at the Polyclinic at Ho, in the Volta Region of Ghana. These cases were diagnosed in the late or advanced stages which makes the prognosis poor. These undiagnosed cases serve as the reservoir for the transmission of the disease in the community. As part of efforts towards zero Leprosy, Ghana has adopted the WHO Global Leprosy Strategy which aims to eradicate Leprosy by 2030. Early detection and contact tracing are at the forefront of Ghana’s action plan following the cases that were reported (16).

Contact tracing is key when cases of Leprosy are reported. Ghana’s reports of Leprosy cases with G2D are telling of the gaps in early detection which also affects contact tracing. The WHO regards high G2D rates as an indication that measures need to be taken especially concerning early detection. If the current situation is left unattended, Ghana faces the challenge of retrogressing in its fight against Leprosy. The study aimed to explore the factors affecting contact tracing for the early detection of Leprosy cases at Ho, Volta Region.

### Theoretical Underpinning

This study is guided by the Centers for Diseases and Prevention adaptation of the Social Ecological Model by Urie Bronfenbenner. The theory explains how various levels of influence (interpersonal, organizational, community, and policy factors) shape human behaviour and health outcomes. (17)

Under the policy-level factors, factors such as interventions, education and institutional policies can be considered. Literature shows that implementing policies that raise awareness of Leprosy is crucial to control cases (18). Adoption of feasible interventions and policies will directly lead to early detection and efficient contact tracing of Leprosy cases.

These interventions also directly affect community-level factors such as stigma. The education and other interventions at the policy level will shape community behaviours and knowledge about Leprosy which will play a significant role in reducing the stigma and misconceptions concerning Leprosy. The misconceptions and stereotypes held by society or a given community lead to the stigmatization of people cured of Leprosy (19). The fear of stigma from the community has also led to delayed case reporting among some people affected by Leprosy. Contact tracing is also affected by stigma as individuals may not want to be stigmatized in the community as a result of being contacts of index cases. Again, beliefs held by the community such as Leprosy being caused by supernatural forces may impede contact tracing for early detection. The reverse is also true where community-level factors can also influence interventions and policies. An effective intervention is tailored to address the issues or needs of the community to which the intervention will be rolled out. Hence, a community’s level of awareness of Leprosy can influence policies on awareness creation.

Individual factors and interpersonal factors are closely related. The individual factors consider a person’s age, educational level, socio-economic status, knowledge of the disease as well as perceived stigma. These factors have a strong linkage with the early detection of cases. This is because factors such as the individual’s knowledge of the disease as well as the ability to recognise symptoms informs a person’s health-seeking behaviour. The interpersonal factors of friends and family are very crucial to contact tracing once a case of Leprosy is detected. Close contacts and household members of an index case have a chance of developing Leprosy if post-exposure prophylaxis is not administered in time (13,20).

## Methods and materials

### Ethical Approval

The proposal for this study was reviewed and approved by the Ghana Health Service Review Committee (GHS-ERC: 064/05/23). All participants signed an informed consent before participation.

### Study Area

The study was conducted at the Ho Polyclinic and at Godokpe, a village at Ho, in the Volta Region of Ghana. The Ho Polyclinic is a Ghana Health Service facility in Ho. The facility provides a wide range of services, but it is known to provide one of the best services for skin conditions. The Polyclinic was established in 1926 as a Leprosy clinic and was converted to a Polyclinic in 1996. Owing to this, the Polyclinic has expertise in Leprosy treatment. The Polyclinic also serves as the main clinic in the southern belt of Ghana that administers treatment for Leprosy. According to the District Health Information Management System (DHIMS) and other records from the Ho Polyclinic, there have been 27 cases of Leprosy recorded in the past year. Godokpe is among the communities in Ho with a number of the cases.

### Study Participants

The population for this study were in three (3) different groups. The first group was PAL who were either on the MDT at the Ho Polyclinic or had completed treatment. The second group was health workers at the Ho Polyclinic and the last group was community members of Godokpe.

### Sample Size Determination and Sampling of Participants

The researcher engaged 6 PAL as well as 6 health workers in reference to findings that saturation of 70-92% will be achieved after conducting 6-12 interviews (21). Maximum variation purposive sampling technique was employed to ensure that all stakeholders who have a role to play in the contact tracing for the early detection of Leprosy were represented. Three (3) focus group discussions were conducted in reference to findings that saturation of 80% will be achieved after conducting 2-3 FGDs (22). Each focus group comprised eight (8) discussants. Community entry was done to know the stakeholders and gatekeepers of the selected community to aid with the purposive sampling of community members.

### Data Collection and Procedure

Data was collected using in-depth interviews guided by a semi-structured interview guide as well as conducting FGDs for community members. With the permission of participants, interviews were recorded and later transcribed verbatim. Notes were taken as a backup. Field assistants were employed to conduct interviews in the local dialect when necessary. Interviews for PAL and the health workers were conducted at Ho Polyclinic. Interview sessions lasted between 30-60 minutes.

Focus group discussions were moderated by trained research assistants since they were conducted in the local language. The discussions took place at Godokpe near the cured Leper’s village. Discussants were made to sit in horseshoe fashion with the moderators and notetakers sitting in front of them. During the discussions, each participant was made to share his or her views on a question posed before proceeding to the next question. An inductive probing strategy was employed to clarify any point raised during the discussion. After the discussion, the moderator summarised the key points as a way of member checking. The FGD lasted for 60-90 minutes.

### Data Analysis

The interviews were translated and transcribed into English. The data was analyzed using thematic analysis. Thematic Analysis is a method of identifying, analyzing and reporting patterns (themes) within data (23). The themes describe salient information which is in line with the research questions. The NVivo version 14 software was used to organize the data. All transcripts were read thoroughly, and themes were identified to answer the research questions. A codebook was developed according to the study objectives and data collected from the field (hybrid inductive-deductive approach). All the transcripts were imported into NVivo. Each transcript was opened and meaningful statements made by various participants were coded into already-developed codes. The transcripts were first coded as free nodes. However, as the coding progressed, the relationship between codes became clearer and thus was transformed into tree nodes. Audit trials were kept and reviewed by the research supervisor for confirmation. Coded segments were regrouped to form main and sub-themes. The results were presented in themes and supported by the most telling illustrative quotes from participants.

## Results

### Background Characteristics of Participants

Overall, 12 in-depth interviews and three (3) FGDs were conducted. A total of 36 participants were recruited for the study. This comprised six (6) health workers (3 males and 3 females), six (6) PAL (3 males and 3 females) and 24 community members (7 males and 17 females). The ages of the participants ranged from 18 to 81 years old. The most frequently mentioned educational attainment was the Basic Education Certificate Examination (19 out of 36) (Table 1).

**Table 1:**
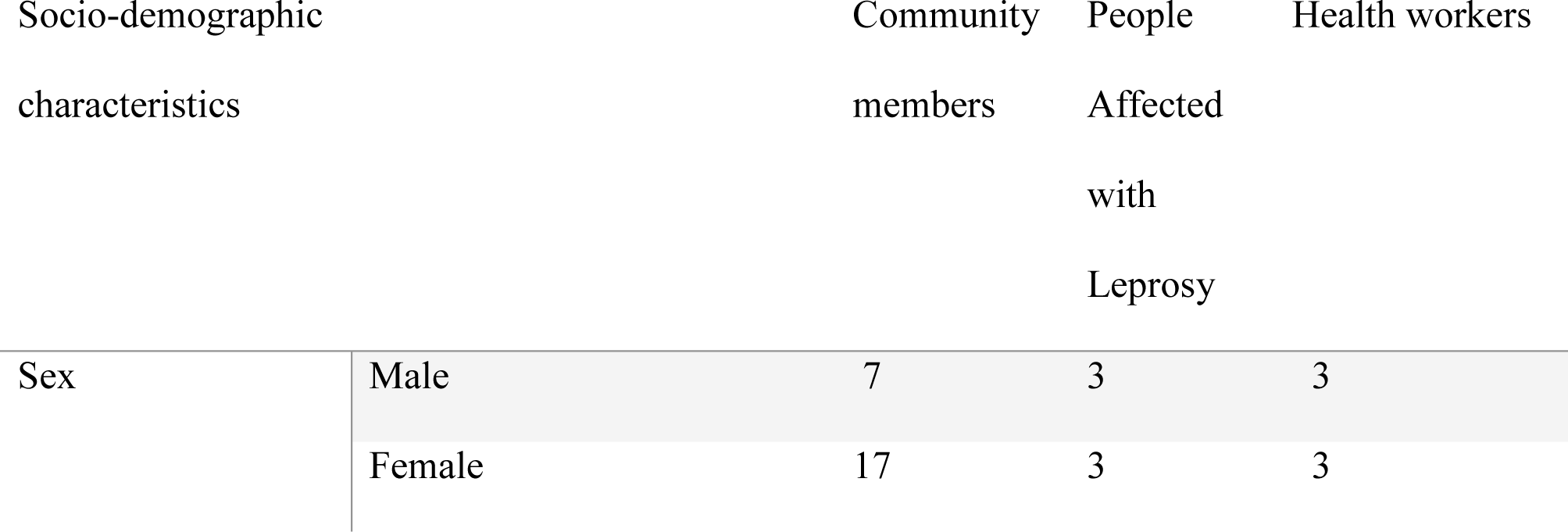
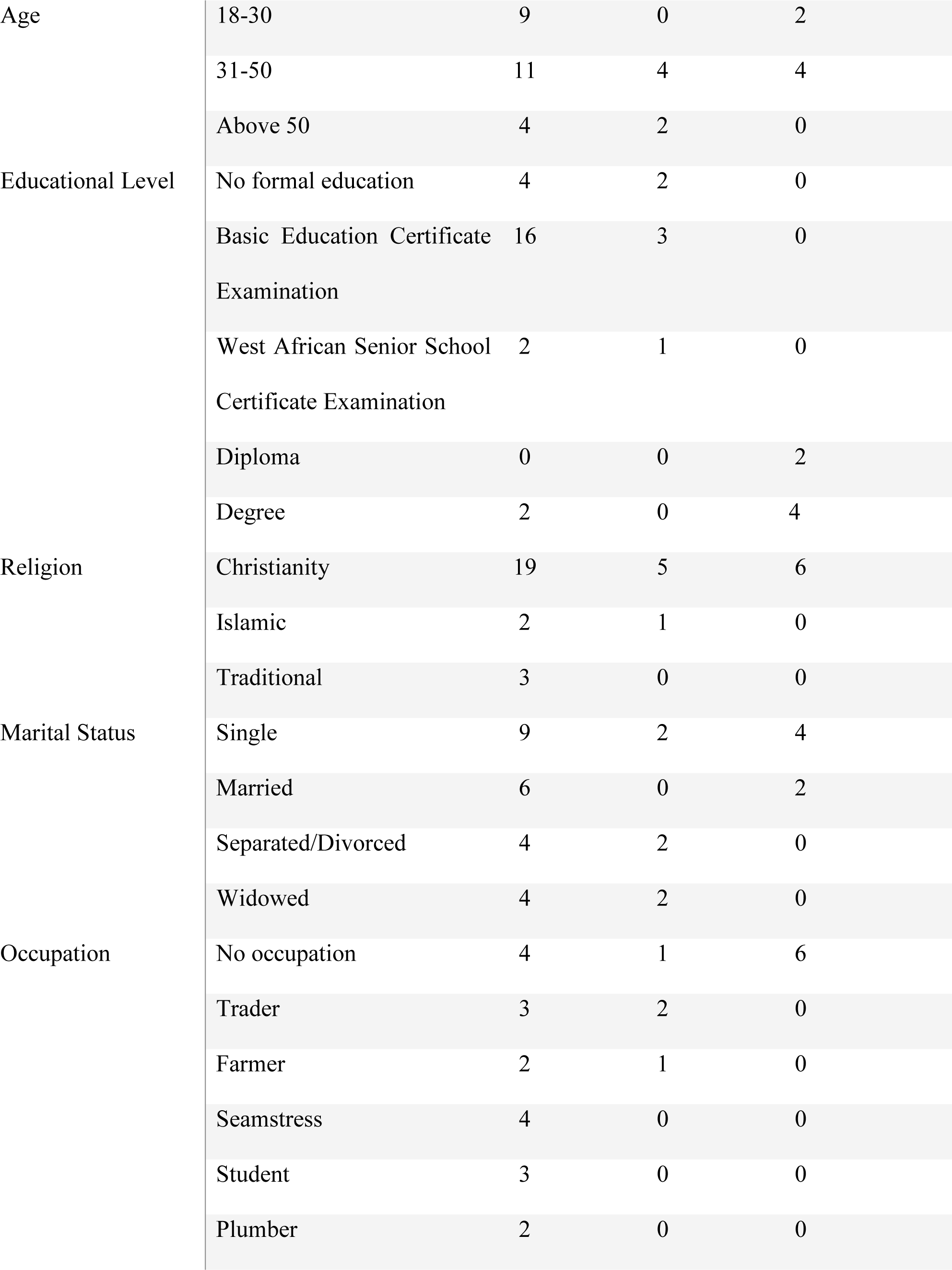
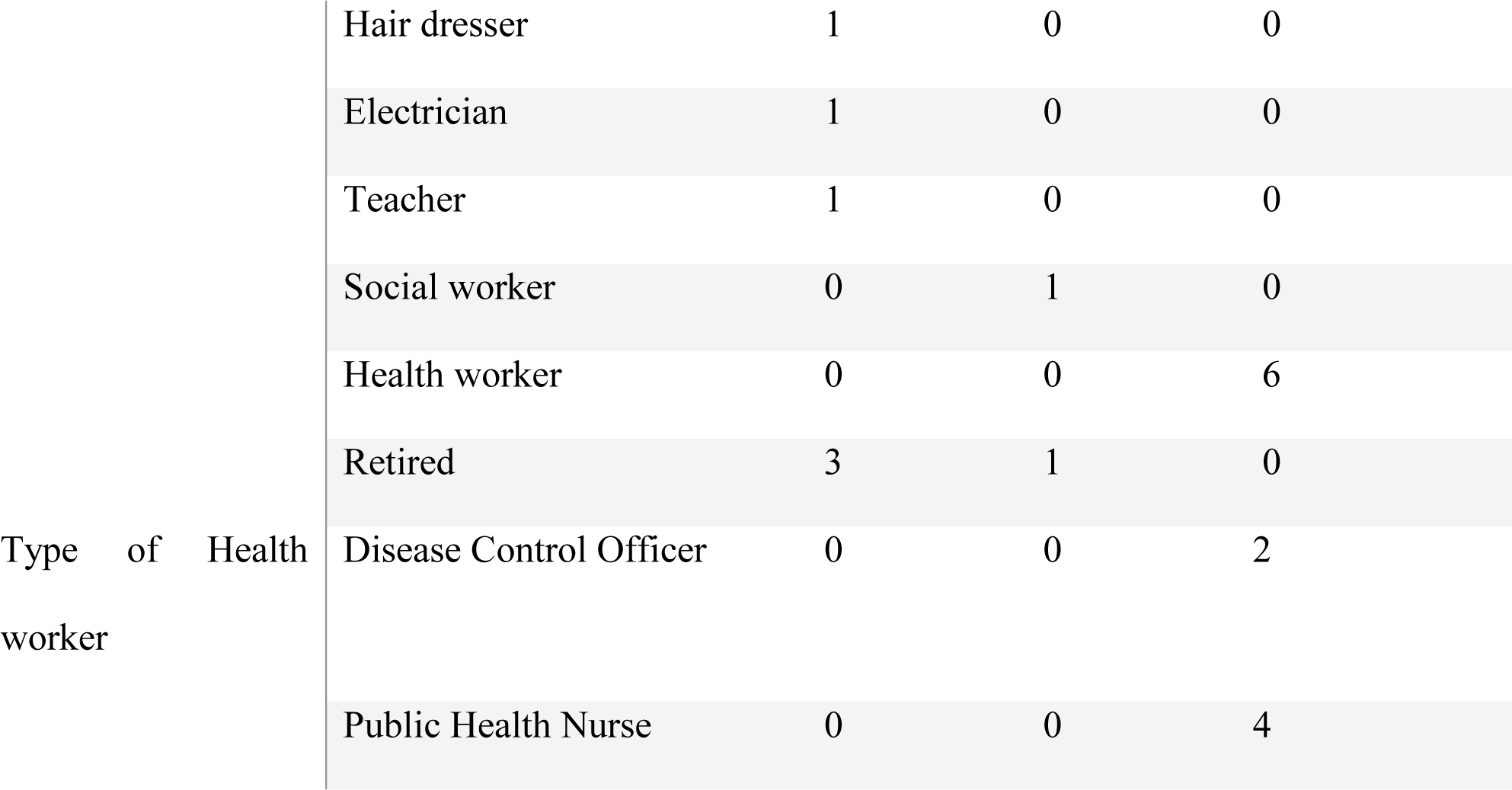
Background Characteristics of Participants.

**Table 2:** Themes from the study.

| Main Themes | Sub-themes |
| --- | --- |
| Facilitators of Early Case Detection of Leprosy | • Knowledge of the symptoms of Leprosy<br>• Understanding of early detection<br>• Awareness creation |
| Barriers to Early Case Detection of Leprosy | • Stigma of the condition<br>• Belief in divine forms of healing and herbal concoctions<br>• Inability to recognize Leprosy symptoms |
| Factors affecting Contact Tracing | • Understanding of contact tracing<br>• The belief that Leprosy is not transmissible |
|  | • Misconception about the cause of Leprosy<br>• Stigma of the condition<br>• Inadequate funds for contact tracing<br>• Knowledge of the Single Dose Rifampicin<br>• Willingness to take the Single Dose Rifampicin<br>• Education on contact tracing |

### Facilitators of Early Case Detection of Leprosy

Participants identified knowledge of the symptoms of Leprosy, understanding of early detection and awareness creation as the facilitators of early detection. They stated that the disease is recognized through abnormal reddish patches on the skin, loss of sensation, skin rashes, loss of sight as well as the loss of fingers and toes. A participant described the condition as follows:

> “Because we are all different, the symptoms vary. Mine started as rashes from mosquito bites. I went to the hospital, I was injected and the rashes disappeared but later reoccurred. I went back to the hospital when they detected that I had Leprosy and asked me to visit the hospital every 8 days for medications.” **PAL_Female**

The health workers also shed light on common symptoms that PAL present prior to diagnosis. A health worker narrated that some of the symptoms presented by PAL are patches, rashes and complaints of loss of sensation:

> “Usually they present with patches and then maybe rashes and sometimes too you see some blisters that look like chicken pox but it’s larger than chicken pox. Some people will also complain that they are not able to feel certain parts of the body which means that their nerves have been damaged.” **Health worker_Male**

Participants were asked to share their views on what early detection is. Participants agreed on the fact that early detection of Leprosy cases is when a person reports to the health facility when the symptoms of Leprosy begin to show. The symptoms that were referred to were usually patches and skin rashes. An illustrative example of such an opinion is described in the following statement:

> “To my understanding, early case detection is when the symptoms are recognized early which helps in diagnosing and treatment of Leprosy as early as possible. For instance, when you see the rashes or patches are not going after a while, you must go to the hospital.” **Community member_Female**

Similarly, one of the PAL expressed that early detection is when a person recognizes changes in the skin and seeks health care for diagnosis.

> “okay to my understanding, early detection is when a person sees any changes on their skin, let’s say rashes that are not going and reports to the clinic to see what is wrong.” **PAL_Male**

All participants were of the view that early case detection of Leprosy is important. To facilitate this, education and awareness creation on Leprosy was considered a sure way. This was the view of one of the healthworkers,

“I think education should be the main focus. Community education; go to the communities and talk to them that in case anyone sees some signs, they should report to the hospital for diagnosis. We can use community durbars, maybe funerals” **Health worker_Female**

### Barriers to Early Case Detection of Leprosy

Participants identified stigma, belief in divine forms of healing and herbal concoctions as well as an inability to recognize symptoms of Leprosy as barriers to early case detection of Leprosy. It was reported by the participants that PAL are mostly shamed and isolated and hence they delay seeking treatment. A community member stated in this regard:

> “From where I grew up, people who suffer the disease are mostly shamed and isolated from people in the community and this prevents them from seeking early treatment.” **Community member_Female**

For one gentleman affected with Leprosy, there was stigma from the community. His family members also distanced themselves from him after hearing of his diagnosis. He narrated as follows:

> “When my sores started becoming a lot, I no longer felt comfortable going into the community because people used to stare at me and did not want to come close. I live alone but no family member wanted to visit me when I told them I was sick with the disease.”

### PAL_Male

The health workers also reported that the stigma associated with Leprosy hinders early detection. A health worker narrated that some of the PAL have the notion that people will run away from them if they get to know that they are carriers of the disease and so prefer to stay at home till the symptoms become worse in the late stages.

> “…some people also do not report because of stigma. Some people say if they go [visit the clinic] and people know that they have the disease, people will start running away from them so they stay at home till it gets to the late stages before they report.” **Health worker_Male**

The study findings showed that there was a tendency for people to resort to divine forms of healing such as the belief that God will heal them. The findings of the study showed that a majority of the participants believe that Leprosy is a spiritual disease and hence seeking medical care may not be the first option. An illustration of this is as follows:

> “Some people say God is the ultimate healer, therefore they may not visit the hospital to get tested and therefore hinder early case detection of Leprosy.” **Community member_Male**

A health worker speaking on the issue also added that Leprosy is believed to be a spiritual illness and hence PAL seek medical assistance when the native doctors and herbal concoctions are unable to relieve them of their symptoms. She expressed herself in these terms:

“Well in this our area some people believe that Leprosy is a spiritual illness or someone has bewitched them so they try other means like going to the native doctors. That is what mostly delays them and they try the herbal concoctions till they realise that they are not healing.”**Health worker_Female**

Some participants were of the view that a person’s inability to recognize the signs and symptoms of Leprosy was a barrier to early detection. A PAL explained that individuals who have ever been affected by the disease are more likely to recognize the symptoms. The implication was that not having someone close to you who has ever had the disease may lead to an inability to recognize the early signs and symptoms leading to late detection.

> “Those who have ever suffered the disease will be able to identify it when they see someone with the symptoms. So in case you don’t have such a person in your house, you wouldn’t be able to identify the disease when someone has the symptoms hence late detection of the Leprosy.” **PAL_Female**

The community members also attested to the fact that ignorance of the disease may be a barrier to early detection. A community member cited ignorance as a barrier to early detection. This view is expressed below:

> “Ignorance of the disease and not knowing or neglecting the symptoms can hinder early case detection.” **Community member_Male**

On the part of the healthcare system, a health worker opined that gaps in the early detection of Leprosy cases are a result of inadequate active case surveillance. He explained that waiting for cases to be reported at health facilities may lead to a delay in the detection of Leprosy cases.

> “It [late detection] could be due to the fact that we are not doing enough active surveillance, we sit down and wait for the cases to come before we diagnose then we will not get the cases on time.” **Health worker_Male**

Another health worker shared his views on the issue saying that going into the community is a sure way of getting Leprosy cases in the early stages. This is illustrated in the quote below:

> “You know, people do not really pay attention to Leprosy nowadays so they do not recognise the symptoms early. Ideally, we should go into the communities to check for some of these symptoms” **Health worker_Male**

The health workers mentioned that there is no test for contacts of index cases of Leprosy and this hinders early detection. They reported that testing for Leprosy at the Polyclinic is done by identifying patches and other changes on the skin as well as checking for loss of sensation. A health worker said in this regard:

> “Over here what we do is we do a physical examination of the person. If there are patches we test for loss of sensation by using a pen or pointed object but if there are no patches or symptoms we do not do any other tests.” **Health worker_Female**

### Factors affecting Contact Tracing

Understanding of contact tracing, beliefs that Leprosy is not transmissible, misconceptions about the cause of leprosy, stigma, inadequate funds for contact tracing, knowledge of the Single Dose Rifampicin, willingness to take the Single Dose Rifampicin and education on contact tracing were the sub-themes that emerged to explain the factors affecting contact tracing.

Participants were asked to explain what they think contact tracing is. The health workers explained what contact tracing is in relation to Leprosy and also gave information on how contacts are traced. Contact tracing was explained as identifying close family, friends or anyone who has been in contact with an index case of Leprosy for a prolonged amount of time. At the Ho Polyclinic, when a case of Leprosy is diagnosed, details of contacts are taken and then a follow-up is done. A health worker explained the process:

> “Yes, so when we get a positive case we have to do a follow-up to prevent the disease from spreading. So when they are diagnosed, they fill out a case-based form. In the case-based form, there is a question on how many people the patient staying with. So for instance the person can say he is staying with 5 people, and then you ask, are you all living in the same house? So we take the client’s number and then fix a date when we can come and visit the house. So everyone that you are staying with or everyone close to you, you bring their names and then we conduct a physical assessment for them too. For all you know, some of them may be having the same condition and we put them on treatment when we get some cases.” **Health worker_Male**

Community members and PAL had a fair knowledge of what contact tracing is. They explained contact tracing as identifying close contacts to index cases. They were however not certain of the benefits. A PAL likened contact tracing for COVID-19 to contact tracing for Leprosy saying that it helps in identifying close family members and friends. However, she did not know why contact tracing should be done for Leprosy. She narrated:

> “Just like COVID, we know that contact tracing helps to identify close family and friends of the person who has the disease but for Leprosy I don’t know why there should be contact tracing.”**PAL_Female**

A participant was of the view that contact tracing is not necessary because the disease is not infectious.

> “Contact tracing is not necessary to me as a person. We have been around people with the disease and we are okay.” **Community member_Male**

The majority of the community members held a strong belief that Leprosy is not an infectious disease and hence cannot be transmitted from one person to another. A community member mentioned that Leprosy is caused by Black magic and hence cannot be transmitted from one person to another.

> “This particular disease, if we want to talk about it we won’t finish today. It is mostly gotten through black magic power and so cannot be transferred from one person to another.” **Community member_Male**

Another community member narrated that both of her parents were people affected by Leprosy. However, she and her siblings do not have Leprosy and hence she is of the view that the disease is not transmissible.

> “It is not transmissible because myself and my siblings were products of two lepers but none of us has suffered the disease.” **Community member_Female**

Others were of the view that Leprosy is caused by supernatural forces. In the study, the majority of the community members and some of the PAL believed that Leprosy is caused by Black magic or is a curse on someone. One of the PAL narrated that she stepped into Black magic powder that was spread in front of her door and got the disease. She explained in the extract below:

> “Mine came as a result of stepping into black magic powder which was spread in front of my door. Initially, I did not know the cause but it is not in my family so I started going for prayers and the pastor told me the cause in his revelation.” **PAL_Female**

The majority of the community members who participated in the FGDs were of the view that Leprosy is caused by supernatural forces. A community member explained that her husband started experiencing the symptoms of Leprosy and was diagnosed with the disease after he stepped on Black magic powder. She narrated the experience as follows:

> “I will say it is spiritual because someone poured black magic powder in our house and when my husband stepped on it, he started experiencing some big rashes on his body which later became sore and when we took him to the hospital, he was diagnosed with Leprosy.” **Community member_Female**

Participants of the study also mentioned that the stigma associated with Leprosy hinders interventions to fight the disease. The community members were of the view that it is considered shameful for people to know that a member of one’s family has Leprosy and hence some of them admitted that they may not be open to being traced as contacts. A participant highlighted the fact that the stigma associated with Leprosy may prevent people from marrying from one’s family.

> “The family traced will be stigmatized. People might not like to marry from such a family.”

### Community member_Female

To add to this, a health worker expressed that the stigma associated with Leprosy causes some family members of the affected to shy away from contact tracing. She further highlighted that the stigma may even rub off on health workers.

> “During the contact tracing some of them [family members] are scared that they will be tagged in the community. At times they ran away, they would not even allow you to speak with them. They don’t avail themselves. Some can be very difficult. Sometimes you the health worker can even be stigmatized [Laughs], they can start calling you Leprosy woman.” **Health worker_Female**

A major barrier to contact tracing reported by the health workers was the inadequacy of funds for contact tracing. A health worker explained that there are Leprosy programmes that should receive regular funding but that does not happen. He also suggested that monetary incentives can be given to health workers to motivate them to trace contacts. He expressed himself as follows:

> “Oh it [challenges with contact tracing] can be addressed but there should be funding. You see, like the way we have TB programmes, we have Leprosy programmes as well. I also know that funds do not come regularly but at times there are donor funds that support such conditions so assuming there is a programme or some funds somewhere, managers can release the funds for contact tracing. And now there is a system that they put in place for like TB for instance to get more cases. The system is that they give staff some package if you can detect two or more TB cases or even when you pick the sample, they give I think 30 cedis or 50 cedis if you report a case, Oti region for example have started that. For Guinea worm, it was 200 cedis initially because they wanted to eradicate it. They can do the same for Leprosy.” **Health worker_Male**

Participants were asked about their knowledge of the Single Dose Rifampicin which is a prophylaxis given to contacts. The majority of the community members and PAL had no idea of the prophylaxis. This is illustrated by the quote below:

> “I have never heard of anything like that [SDR].” **PAL_Male**

The community members also expressed a lack of awareness of the SDR. One person said:

> “I do not know what that medicine is. I have never heard of it”. **Community member_Male**

However, a few community members and PAL had knowledge of SDR. A community member mentioned that the SDR is given to contacts to prevent them from getting the disease. He expressed himself as follows:

> “They give the medicine to the contacts to prevent them from contracting the disease.”

### Community member_Male

One PAL mentioned that she is aware that prophylaxis is given to close contacts who have not been diagnosed with Leprosy.

> “I know the prophylaxis is given to those who don’t have the disease but are close to someone who is infected.” **PAL_Female**

A health worker confirmed that though he knows of the prophylaxis, it is not administered to contacts. He explained that there are occasional delays in the supply of MDT (Multidrug Therapy) due to shortages. Owing to this, the Rifampicin which is part of MDT is not enough to be given to contacts as well.

> “Yes, I know the Rifampicin can be given as prophylaxis for the contacts, but we do not give that here. You know, as I mentioned sometimes there are delays in the supply of the MDT so already we a short of the Rifampicin for the cases how much more the contacts.” **Health worker_Male**

The community members were asked about their willingness to take SDR if traced as contacts. The majority of the community members expressed unwillingness to take the prophylaxis. A community member was of the view that prophylaxis may trigger other diseases in him so he will not take the drug if he does not have Leprosy. He expressed himself by saying:

> “I will not accept the medication if I test negative for the disease because it might trigger another disease in me.” **Community member_Male**

Inadequate knowledge about prophylaxis was a reason given by one of the community members who expressed unwillingness to take prophylaxis.

> “I don’t know much about the medication so if there is nothing wrong with me, I will not take the medication.” **Community member_Female**

Contrary to these views, a few community members expressed willingness to take prophylaxis if the disease runs in the family to curb the spread.

> “Since it is a disease and we wouldn’t want it to spread among us as a family, I will gladly accept it.” **Community member_Female**

The findings of the study indicate that education or awareness creation is the way to go to facilitate contact tracing. A participant was of the view that people should be educated to know what exactly contact tracing is to increase its acceptability.

> “I think when one person from a family has the disease, we should all avail ourselves as a family, friends and neighbours for testing to be sure we also do not have the disease. They must be educated to know what exactly is contact tracing.” **Community member_Female**

## Discussion

### Facilitators of Early Detection

The findings of the study showed that even though community members and PAL had some knowledge of Leprosy and its symptoms, the information was diluted with some misconceptions. Owing to this, participants reported that some symptoms of Leprosy go unrecognized or are trivialized resulting in late detection. Adequate awareness of Leprosy and its symptoms was found to facilitate early detection of Leprosy. Consistent with the literature, the symptoms of Leprosy which include unaesthetic hyperpigmented patches of the skin, deformities, blindness, and damage to the peripheral nerves were known and mentioned by the participants (1,3,6,24). Though community members and PAL in the current study demonstrated some knowledge of the symptoms of Leprosy, they acknowledged that their knowledge may be scant and hence education and awareness creation are needed. Findings from a study in India revealed that increasing awareness of the early signs of Leprosy was the most effective way to increase early case detection as compared to contact tracing and the administration of post-exposure prophylaxis (18). This study was conducted in India, but the current study shows that the situation in Ghana is not sui generis. None of the participants mentioned contact tracing or prophylaxis as a means of facilitating early detection of Leprosy. Furthermore, the PAL who were interviewed reported that they had no suspicion of being infected with Leprosy despite the symptoms they had before diagnosis. This highlights the need for awareness creation on the early signs of Leprosy.

An understanding of what early detection of Leprosy is was shown to be crucial to facilitating early detection. The views on what early detection of Leprosy means were varied in the current study. Some participants were of the view that early detection is when a person seeks medical health before symptoms progress to disabilities. Others were of the view that early detection occurs when a person first notices any unusual changes in the skin and seeks medical attention. A specific timeline for early detection was not reported by participants. This is because the participants held the view that the prognosis for Leprosy differs per person. Similarly, there are different views on what the period of early detection of Leprosy should be in the literature. A threshold of 6 or 12 months has been proposed in literature as a uniform definition for detection delay in Leprosy (19). However, the WHO purports that on average, a case of Leprosy is considered to be reported early when the duration between the onset of the disease and the diagnosis is less than 2 years or when there is no G2D (25). The findings of the present study revealed that some individuals who were diagnosed within these time frames had started developing G2D. If the development of G2D is a marker of late detection, then the time frames reported in the literature for early detection are questionable as individuals may develop disabilities at any time. An understanding of early detection and its benefits is also key in helping individuals appreciate the importance of seeking medical assistance when any unusual changes on the skin are noticed.

### Barriers to early detection

The stigma associated with the disease was suggested as a possible reason why people affected by the disease may delay seeking treatment. A study in Ghana revealed that the stigma and discrimination from family, friends and community members even after a person is cured of Leprosy leaves the individual with no choice but to seek refuge and isolate in the Leprosarium (19). The findings of the current also echo the results of a study conducted in Western Nigeria (6). The findings were that the perceived supernatural causation of Leprosy coupled with the stigma associated with the disease frustrates early detection (6).

Another barrier to early detection mentioned by the participants is the reliance on God and traditional forms of medicine for healing. This is most likely because the disease is believed to be caused by supernatural forces. The use of herbal concoctions often delays early detection and leads to an aggravation of the symptoms. According to a study in Ethiopia, the use of traditional forms of healing which proves ineffective over time is a major reason for late detection (26). However, the present study reported that some PAL believe in both biomedical and alternate forms of treatment. This report is consistent was the findings in Nepal on the challenges of health-seeking behaviour among PAL (27).

Ignorance of Leprosy and its signs and symptoms hinders early detection. The participants of the current study reported that early signs of Leprosy such as rashes are trivialized and believed to be skin conditions that will disappear with time. The link between symptom misinterpretation and delays in diagnosis is further emphasized in the literature (28,29).

The inability to test asymptomatic carriers also posed a barrier to early detection of Leprosy. Clinical diagnosis of Leprosy is difficult in the early days of infection as well as among asymptomatic carriers (30). Owing to this, diagnosis of Leprosy is done when an individual starts showing symptoms of the disease. Like in the literature, an accurate test for early detection of asymptomatic individuals affected with Leprosy is yet to be developed despite the numerous attempts to develop one (31). This has implications for early case detection.

### Factors Affecting Contact Tracing for Leprosy

Participants of the study were asked about their understanding of what contact tracing is in relation to Leprosy. The general understanding of contact tracing was that it is the process of identifying family, close ties or individuals who have been in contact with an index case of Leprosy within a given period. The main rationale behind contact tracing which is to identify carriers of the disease early and control the spread of the infection was reported by the participants. However, there were gaps in the implementation of effective contact tracing.

The issue of stigma was also mentioned as a barrier to contact tracing. As reported in the literature, due to the stigma associated with the disease, some family members and friends shy away from PAL (19,32,33). These family members and friends are the contacts of this individual and hence if they do not want to be associated with the PAL, then contact tracing is compromised.

The misconception that Leprosy is not an infectious disease among the majority of the community members and PAL is a barrier to contact tracing. A participant also expressed the view that contact tracing is not important. This may be because it is believed that Leprosy is caused by supernatural forces. This belief coupled with the belief that Leprosy cannot be transmitted makes it difficult for people to appreciate the importance of contact tracing. It has been established that though the causative organism for Leprosy is *Mycobacterium leprae*, the mode of transmission is unknown (2,34). Contrary to this, there was no mention of Leprosy being a bacterial infection among the community members and the PAL. The commonly reported cause of Leprosy was witchcraft or Black magic. This has implications for the spread of the disease as well as interventions for contact tracing. It is however known that there is an association between the presence of the BCG scar on an individual and an increased immunity to the infection It has been inferred that the BCG vaccination is to some extent efficacious in preventing Leprosy according to the findings of a study in the Sene District in the Bono East Region (35). The indication that a person has been vaccinated is the presence of a scar on the upper right arm. Ghana introduced routine BCG vaccination into its Expanded Programme on Immunization in 1978. Given that a number of the participants in the current study may have been vaccinated, immunity to the infection may have increased hence the belief that Leprosy is not infectious as some participants testified to being in close contact with index cases and not exhibiting any symptoms. It is also known that Leprosy has a lengthy incubation period of up to 20 years without symptoms (4). This implies that an individual may be in continuous contact with a PAL without exhibiting symptoms and hence believe that it is not infectious only to start exhibiting symptoms years after.

A key barrier to contact tracing mentioned by the health workers was financial constraints. The Polyclinic serves as the main clinic in the southern belt of Ghana that administers treatment for Leprosy. There is therefore a challenge with following up to do contact tracing for individuals who do not live in the Ho vicinity. Again, the health workers reported that inadequacy in active surveillance is a barrier to contact tracing. This was also attributed to the inadequacy of funds. Passive surveillance where there is a reliance on PAL to report at health facilities is a poor way to fight the condition (36).

Studies show that the SDR is efficacious in preventing Leprosy in close contacts. This was tested in a study on preventing Leprosy with retrospective active case finding combined with SDR for contacts in an area in Cambodia (20). To the best of the researcher’s knowledge, similar studies have not been replicated in Ghana. However, all the health workers interviewed had knowledge of SDR and its efficacy. However, they admitted that it was not available to contacts. They further highlighted the challenge of getting the MDT for PAL. The Rifampicin which is supposed to be administered to contacts is a component of the MDT and hence the challenge with the supply of MDT affects the supply of the Rifampicin to contacts.

Again, knowledge of SDR among community members and PAL was low. Willingness to accept the SDR was also low though some community members and PAL expressed willingness to take the prophylaxis. Contrary to this, a study on the Leprosy control programme implemented in Tanzania, Brazil, Myanmar, Nepal, India, Indonesia and Sri Lanka showed that the prophylactic treatment was generally accepted among contacts (37). This low acceptability of SDR in the current study can be attributed to the fact that the knowledge of the prophylaxis is low. Again, the notion that Leprosy is not transmissible can affect an individual’s willingness to take prophylaxis.

To facilitate effective contact tracing from the viewpoint of the health workers, there should be adequate allocation of funds. Little to no allocation of funds to neglected tropical diseases raises the concern of the re-emergence of these infectious diseases. Increasing awareness was also mentioned as a facilitator of contact tracing. Increased awareness will make people appreciate the importance of contact tracing.

### Study Limitation

The language barrier encountered during data collection and transcription is the main limitation of this study. This limited the researcher’s ability to verify the quality of the data collected. However, the researcher ensured that field assistants were well-versed with the interview guides to ensure that quality data was collected. Regarding the transcription, the researcher employed individuals aside from the field assistants who did the initial transcription to do a back-to-back transcription to ensure that little to no data was lost.

## Conclusion

Leprosy was perceived as a supernatural condition. This perception affected the health-seeking behaviour of PAL inclining them towards spiritual remedies. This resulted in a delay in case detection as these spiritual health outlets do not have the capacity to diagnose and treat such conditions. The belief that the condition was not infectious is a community-level factor that affected the early seeking of treatment which affected case detection. The lack of funds was a major issue mentioned by the health workers. Aside from this, stigma and low awareness of the importance of contact tracing were also mentioned as barriers to contact tracing. The study also revealed that there was low awareness and willingness to take the Single Dose Rifampicin. Overall, the study results indicate that raising awareness of contact tracing and early detection will address the majority of the challenges reported.

## Data Availability

All relevant data are within the manuscript and its Supporting Information files

## Acknowledgements

The authors wish to thank all the study participants for their time.

